# Peripheral blood cytokines as markers of longitudinal recovery in white matter microstructure following inpatient treatment for opioid use disorders

**DOI:** 10.1101/2024.10.09.24315171

**Authors:** Eduardo R Butelman, Yuefeng Huang, Sarah G. King, Pierre-Olivier Gaudreault, Ahmet Ceceli, Greg Kronberg, Flurin Cathomas, Panos Roussos, Scott J. Russo, Rita Z. Goldstein, Nelly Alia-Klein

## Abstract

**Background:** Heroin and other opioid use disorders (HUD and OUD) cause massive public health morbidity and mortality. Although standard-of-care medication assisted treatment (MAT) exists, little is known about potential predictors of change during recovery. Recovery can include normalization of the brain’s white matter (WM) microstructure, which is sensitive to cytokine and immune signaling. Here we aimed to determine whether blood-based cytokine/immune markers can predict WM microstructure recovery following medication-assisted treatment.

**Methods:** Inpatient Individuals with HUD (iHUD; n=21) and healthy controls (HC; n=24) underwent magnetic resonance scans with diffusion tensor imaging (DTI) and provided ratings of drug cue-induced craving, arousal and valence twice, earlier in treatment and ≈14 weeks of inpatient MAT (with methadone or buprenorphine) thereafter. At this second session (MRI2), they also provided a peripheral blood sample for multiplex relative quantification of serum cytokine/immune proteins (with a proximity extension assay, Olink). We explored the correlation of a multi-target cytokine biomarker score (based on principal component analysis of 19 proteins that differed significantly between iHUD and HC) with change in whole-brain DTI (ΔDTI; MRI2 - MRI1) metrics (fractional anisotropy, mean diffusivity, and axial diffusivity) across the 14 weeks of MAT.

**Results:** The cytokine biomarker score, obtained at the MRI2 stage, was correlated with ΔDTI metrics in frontal, fronto-parietal, and cortico-limbic WM tracts (e.g., including the genu of the corpus callosum, anterior corona radiata, and others). In a follow-up analysis, specific cytokines represented in the multi-target biomarker score, such as the interleukin oncostatin M (OSM), colony stimulating factor (CSF21), and the chemokine CCL7 were correlated with similar ΔDTI metrics in iHUD, but not in HC. Levels of other specific cytokines (i.e., CCL19 and CCL2) were negatively correlated with change in cue-induced craving or arousal. Thus, lower levels of the aforementioned cytokines were correlated with an increase in cue-induced craving or arousal across the two stages (MRI2 - MRI1).

**Conclusions:** Studied as a multi-target biomarker score, or as individual targets, peripheral serum cytokines are highly accessible biomarkers of WM microstructure recovery in iHUD undergoing inpatient MAT.

## Introduction

Chronic exposure to mu-opioid receptor (MOR) agonists such as in heroin (HUD) and other opioid use disorders (OUD) cause major morbidity and mortality (1,2), constituting a national epidemic. While medication-assisted treatment (MAT; e.g., with methadone or buprenorphine) is the effective standard of care, many individuals with HUD (iHUD) either do not receive treatment, drop out, or relapse (3,4). Limited knowledge about potential biomarkers of treatment outcomes (inclusive of recovery) of persons undergoing MAT severely limits the development and deployment of timely intervention and prevention efforts. Hence, there is a critical yet unmet need for the identification of new and accessible brain-based biomarkers of risk and resilience in iHUD during treatment.

Among the molecular targets dysregulated by chronic MOR agonist exposure (including to heroin) are several cytokine systems, as recently reviewed by our group (5) and others (6,7). Cytokines constitute a pleiotropic group of signaling molecules that act on cognate receptors in peripheral blood mononuclear cells (PBMC) and other leukocytes (8), as well as central glia and neurons (7). The major canonical cytokine families include interleukins (IL), chemokines, growth factors and tumor necrosis-factor (TNF) - related proteins (9), which act in functional signaling networks (9–11); specific leukocyte subsets or cytokines from blood can also affect central neuro-glial processes, by migration through the blood-brain barrier (9,12,13) We recently reported that serum analysis of a multiplex cytokine and inflammatory panel of proteins revealed significant differences in iHUD (n=21) undergoing inpatient MAT versus healthy controls (HC, n=24) (14). Furthermore, in this previous study, a multi-target “cytokine biomarker score” based on principal component analysis (PCA) of 19 specific targets (chemokines, interleukins, growth factors, and TNF-related proteins) robustly differentiated iHUD from HC, adjusting for major demographic and clinical factors. Other studies similarly show changes in blood levels of several cytokines in iHUD compared to HC, and changes in mRNA expression of cytokine genes, both in PBMC and in the brain (5,8). These central and peripheral mechanisms may be broadly conserved, since levels of several cytokines are altered in blood of rats with extended access fentanyl self-administration (15), and there are alterations in expression of cytokine-related genes in the striatum of mice who chronically self-administered oxycodone (e.g., oncostatin M receptor, OSM-R) (7). Importantly, the blood-brain barrier can also become functionally compromised after chronic MOR agonist exposure or withdrawal, potentially exacerbating neuroinflammatory effects (16,17).

The brain’s white matter (WM) is composed of myelinated neuronal tracts (axonal bundles), which underlie functional connectivity networks (18,19). Myelination is an active regulatory process, based on interactions between glia, especially oligodendrocytes and oligodendrocyte progenitor cells (as well microglia and astrocytes) and neurons (20,21), affecting neurotransmission and neuroplasticity (21). Crucially, myelination and neuroinflammation are governed by complex signaling networks, including cytokines and cognate receptors, as well as other proteins (22,23). Of relevance to these processes and HUD, a recent preclinical study showed that MOR-mediated reward depends in part on interactions between neurons and oligodendrocytes (24). Also, there is robust mRNA expression of *OPRM1* (the gene that encodes for MOR) in brain glia (including microglia, oligodendrocytes and oligodendrocyte progenitor cells) of humans post-mortem (25). Indeed, chronic exposure to MOR agonists causes transcriptional and functional changes in molecular targets in CNS neurons (25–27), but also in oligodendrocytes and other glia (25), especially for diverse cytokine and neuroinflammatory genes (25).

White matter microstructure can be examined in humans with diffusion tensor imaging (DTI). We and we and others have documented widespread deficits in WM microstructure in individuals with substance use disorders including HUD (encompassing reduced fractional anisotropy; FA) (28–30) as correlated with enhanced disease severity (e.g., increased craving and longer periods of regular use) (28). Importantly, these WM microstructure impairments appear to be dynamic, recovering in time with treatment. Thus, following inpatient treatment of about 14 weeks, FA increased especially in frontal major callosal projections, association tracts, and the genu and body of the corpus callosum as associated with reduced craving in iHUD (n=30) (31). Another very recent longitudinal study in iHUD (with an 8-month interval, associated with increasing abstinence) also detected an increase in mean FA in some frontostriatal tracts (32).

Here we test whether the WM microstructure recovery we found in iHUD undergoing MAT correlates with readily accessible blood-based cytokine biomarkers. We hypothesized that serum cytokines/immune proteins (from a multiplex assay) will correlate with changes into treatment and abstinence in WM microstructure and drug cue-induced subjective ratings (e.g., of craving, arousal and drug valence).

## Methods

### Participants and Diagnostic procedures

Individuals with HUD (N=21) were recruited from an inpatient treatment facility in New York City, and the HC (N=24) were recruited from the surrounding community. The study was approved by the Institutional Review Board of the Icahn School of Medicine at Mount Sinai; participants gave written informed consent. A highly trained study team supervised by senior clinical psychologists conducted participant evaluations that included the MINI International Neuropsychiatric Interview (33) (version 7.0.0. For DSM-5) and the Addiction Severity Index (34).

### Inclusion criteria for all participants

Age 18-65 and capacity to understand and provide informed consent in English. **Inclusion criteria for iHUD:** Meet DSM-5 criteria for OUD, with heroin as the drug of choice or main reason for treatment. The iHUD were inpatients under MAT on methadone (n=17) or buprenorphine (n=4) daily maintenance. **Exclusion criteria for all participants:** 1) present or past history of DSM-5 diagnosis of psychotic disorder or neurodevelopmental disorder; 2) history of head trauma with loss of consciousness (>30 min); 3) history of neurological disorders including seizures; 4) current use of any medication (with the exception of MAT in the HUD) that may affect neurological functions; 5) current medical illness and/or evident infection including cardiovascular disease (e.g., high blood pressure), as well as metabolic, endocrinological, oncological, or autoimmune diseases, and infectious diseases common in individuals with substance use disorders including Hepatitis B and C or HIV/AIDS. We did not exclude HUD subjects for history of other drug addiction (e.g., alcohol, marijuana, stimulants) or other psychiatric disorders with high rates of comorbidity with drug addiction (e.g., depression, anxiety, post-traumatic stress disorder); 6) MRI contraindications including any metallic implants, pacemaker device, or pregnancy; 7) a positive breathalyzer test for alcohol; and 8) MRI quality assurance, including the presence of incidental findings in the WM as indicated by a radiologist, insufficient quality of diffusion data, or an MRI session that did not include a diffusion sequence. **Exclusion criteria for HC specifically:** Lifetime history of substance use disorders, including for alcohol, or positive urine test for any drug of abuse. The HC were recruited and assessed by the same study team with the same tools, but without completing the addiction-specific scales.

### Behavioral variables

Demographic (age, sex, race, body mass index) and clinical variables (sleep and drug use measures) are included in Table 1. Because depression, anxiety and stress exposure are comorbid with OUD and can be associated with functional change in cytokine mechanisms (13,35,36), the participants completed the Beck Depression Inventory (BDI-II) (37,38), Beck Anxiety Inventory (39) and Perceived Stress Scale (PSS-10) (40). In the iHUD, we also examined methadone/buprenorphine dose (either from documented report, or from self-report) and duration of current heroin abstinence. Age of first heroin use, age of onset of regular use, and number of years of regular use (excluding abstinence periods) were also assessed (41,42) (Table 1). Following each of the two imaging scans (described below) we quantified self-reported ratings of cue-induced craving, valence and arousal in response to drug-related pictures, using a 5-point scale (increasing numbers indicating greater ratings for these measures), as described previously (43).

**Table 1:** Demographic and clinical variables.

| Variable | iHUD<br>(n=21) | HC<br>(n=24) | Group differences;<br>Mann-Whitney U or<br>Fisher's exact test |
| --- | --- | --- | --- |
| Age: mean (95%CI) | 42.0 (38.5-45.5) | 40.7 (36.0-45.4) | NS |
| Sex: M / F | 17M / 4F | 15M / 9F | Fisher's exact test; NS |
| Race: White / Black / Other | 18 / 0 / 3 | 12 / 8 / 4 | Fisher's exact test; p=0.006 |
| Education years: mean (95%CI) | 16.6 (15.3-17.9) | 12.1 (11.2-12.9) | U=45.5; p<0.0001 |
| Body mass index (BMI) at MRI2:<br>mean (95%CI) | 32.3 (29.4-35.3) | 26.7 (24.9-28.6) | U=110; p=0.0009 |
| Sleep hours in prior night to MRI2:<br>mean (95%CI) | 6.54 (5.6-7.5) | 6.51 (5.9-7.2) | NS |
| Perceived stress PSS-10 scores at<br>MRI2: mean (95%CI) | 20.5 (17.7-23.3)<br>n=20 | 12.3 (9.5-16.0) | U=104; p=0.001 |
| BDI-II Depression scores at MRI2:<br>mean (95%CI) | 15.5 (9.9-21.2)<br>n=19 | 3.8 (1.8-5.7) | U=78; p=0.0001 |
| BAI Anxiety scores at MRI2: mean<br>(95%CI) | 14.1 (8.8-19.4)<br>n=20 | 2.8 (1.1-4.4) | U=85.5; p=0.0001 |
| Age of onset of first use of heroin:<br>mean (95%CI) | 23.6 years (17.4-27.8)<br>n=20 | N/A | N/A |
| Age of onset of regular use of heroin:<br>mean (95%CI) | 25.1 years (21.2-28.9)<br>n=20 | N/A | N/A |
| Years of regular heroin use:<br>mean (95%CI) | 8.5 years (5.9-11.2)<br>n=19 | N/A | N/A |
| Heroin abstinence duration:<br>mean (95%CI) | 198 days (130-268) | N/A | N/A |
| Methadone daily dose:<br>mean (95%CI) | 91.9 mg (63-121)<br>n=17 | N/A | N/A |
| Years of regular alcohol use* | 12.3 (5.9-18.6)<br>n=17 | 13.2 (6.6-19.8)<br>n=18 | NS |

\*From the Addiction Severity Index

### Cytokine assays

Blood samples were obtained by venipuncture on the day of the second MRI scan (MRI2), or at the nearest practical date (mean interval between MRI2 and blood sample=1.3 days across all participants; n=45). The blood sample was obtained in the time range of 09:00-17:00 hours (i.e., at least 1 hour after the daily MAT administration in the iHUD). Samples were centrifuged (10 minutes; 1,200 G) within ≈1 hour of collection, and serum stored at -80oC. Serum samples were analyzed with the Olink Target 96 Inflammation panel (Olink, Uppsala Sweden), following manufacturer’s instructions (34), by the Human Immune Monitoring Center at the Icahn School of Medicine at Mount Sinai. This validated panel measures 92 targets (including chemokines, ILs, growth factors, TNF-related molecules as well as other inflammation-related proteins; full target list can be found at: https://olink.com/products-services/target/inflammation/). The assay provides relative quantification as normalized protein units (NPX) for each target, on a log2 scale. If a target had ≥50% of samples below the Limit of Detection (<LOD) within either group, the target was not analyzed (35). Of 92 targets in the assay, 14 were excluded for this reason (where ≥50% of the samples were <LOD for: IL2, TSLP, IL22, IL2RB, IL1 alpha, RA1, Beta-NGF, NRTN, and IL5, IL24, IL13, IL20, IL33, IL4, LIF, NRTN). The remaining 78 targets were analyzed (including individual values <LOD) as in prior studies (44). Also, individual outliers (>±3SD from the group mean) were removed from the analyses, and were replaced by imputation for the principal component analysis (see below).

### MRI acquisition and diffusion tensor imaging

All individuals including HC were scanned twice, as part of a larger study, as recently described (31). Briefly, MRI acquisition was performed using a Siemens 3.0 Tesla Skyra scanner (Siemens Healthineers AG, Erlangen, Germany) with a 32-channel head coil. Diffusion echo-planar sequence was acquired with opposite phase encoding along the left-right axis, monopolar diffusion encoding with 128 diffusion-weighted images (2×64 for each encoding phase) at single shell maximum b=1500 s/mm^2^, 13 reference images at b=0 s/mm2, field of view (FOV)=882×1044 mm, repetition time (TR)=3650 ms, echo time (TE)=87 ms, bandwidth=1485 Hz/px, and 80° flip angle, multiband=3, no in-plane acceleration, and 1.8 mm isometric voxel size, with a total acquisition time of approximately 7 minutes. Anatomical T1-weighted structural images were acquired using the following parameters: 3D MPRAGE (Magnetization-Prepared Rapid Gradient-Echo) sequence with FOV of 256 × 256 × 179 mm3, 0.8 mm isotropic resolution, TR/TE/TI = 2400/2.07/1000 ms, 8° flip angle with binomial (1, −1) fat saturation, 240 Hz/pixel bandwidth, 7.6 ms echo spacing, and in-plane acceleration (GRAPPA- generalized autocalibrating partially parallel acquisitions) factor of 2.

### Statistical analyses for phenotypic variables and cue-induced subjective ratings

Table 1 shows the distribution of sex and race across groups, analyzed with Fisher’s exact test. Demographic/clinical data (body mass index, age, sleep hours, depression, anxiety and perceived stress were compared across groups with Mann-Whitney U tests. Cue-induced subjective cravings, arousal and drug valence (all on a 1-5 scale, with increasing numbers denoting greater ratings) were analyzed as Δvalues (i.e., MRI2-MRI1). Thus, a negative Δ value would indicate a decrease for a specific rating in MRI2 vs MRI1. Pearson correlations were carried out between levels of 19 cytokines (see below) and Δdrug cue-induced ratings for craving, valence and arousal (43). The α for significance was set at the p<0.05 level, with a FDR correction for the 19 targets studied.

Normality of variables was examined with the D’Agostino and Pearson test, followed by examination of quantile-quantile plots, if necessary. The overall alpha level of significance was set at the p=0.05 level.

### Quantification of individual cytokines and principal component analysis

Normalized protein units (NPX; log2 scale) were analyzed with Wilcoxon’s rank-sum tests for group differences; p-values were corrected for multiple comparisons with the False Discovery Rate (FDR; 5% cutoff level) approach (45). After identifying 29 targets that showed significant group differences from the previous step (including FDR correction), data from the 19 targets from canonical cytokine families that differed significantly between the iHUD vs. HC (after multiple comparison correction) were entered into a principal component analysis (PCA) for dimensionality reduction (35,46), using z-scores to standardize values. Scores for the first component (PC1) accounted for the greatest proportion of variability (40.9%). Because PC1 requires complete data, individual outliers (i.e., greater than ±3SD within each group mean) were replaced by multiple imputation (missMDA procedure in R) (47). Nineteen principal components were calculated in the algorithm, using a 95% threshold for significance, using 1,000 Monte Carlo simulations (GraphPad Prism). The complete PC1 results on these participants was recently reported (14). The PC1 data were regressed against the whole-brain WM changes (i.e., ΔDTI for the four metrics).

### Analyses of change in WM metrics during treatment

The preprocessing of the diffusion MRI data was computed according to established pipelines with MRtrix3 (48) and FMRIB Software Library (FSL 6.0) toolboxes, as fully described (28,31). Tensor-based quantitative maps of diffusion metrics: fractional anisotropy (FA), mean diffusivity (MD), axial diffusivity (AD), and radial diffusivity (RD) were generated and used to perform Tract-Based Spatial Statistics (TBSS) (30). Whole-brain voxelwise analyses across all participants was conducted by first creating a WM skeleton and registered to the MNI152 standard space. The whole-brain correlation analyses were then performed between the four ΔDTI metrics (i.e., FA, MD, AD, RD) and the cytokine PC1 score (based on the 19 cytokines with significant group differences, Figure S1) to test for slope differences between the iHUD and HC groups. We also tested the correlations between cytokine PC1 score and the four DTI metrics at MRI1 and MRI2 separately, for completeness. Within-group correlations were performed when significant group differences in slopes were detected. Analysis was performed via the FSL tool “Randomise”, a general linear model for non-parametric permutation inferences via 10,000 permutations. Specifically, the ΔDTI metric maps (for whole-brain WM changes for each of the four diffusion metrics) were first computed. Then these individual maps (for the different DTI metrics: FA, MD, AD and RD) were entered into separate models with the mean-centered cytokine PC1 scores. In a follow-up, we repeated the same analyses with normalized protein units (NPX; on a log2 scale) from the individual cytokines from each canonical family with the highest PC1 loadings (i.e., CCL7, OSM, CSF1, TNFRSF9; Figure S1) (14). We also performed correlation analyses between the Δmaps of the four DTI metrics and variables including intracranial volume (estimated by Freesurfer from the individual T1- weighted structural scan), years of education, age, sex, and body mass index, to rule out confounding factors. All DTI analyses accounted for multiple comparisons by using the Threshold-Free Cluster Enhancement (TFCE) correction (49), which enhances areas of signal exhibiting spatial contiguity to better discriminate clusters.

## Results

As shown in Table 1, the groups were well matched on age and sex yet different on some variables. Individuals with HUD had a different racial distribution (more Caucasian and no African-American); they also had a higher body mass index and higher depression, anxiety and perceived stress scores. iHUD also had a shorter number of years of education. The iHUD and HC had a similar number of years of alcohol use.

### Cytokine data

As recently reported for this cohort (14), nineteen cytokines were significantly different between iHUD vs. HC, and these were analyzed with PCA (see also Supplementary Materials, Table S1 and Figure S1).

### Cytokine PC1 scores in correlation with DTI change metrics

We found significant group differences only in correlations between cytokine PC1 scores and the changes in DTI metrics (MRI1 - MRI2) (Figure 1) but not with the absolute DTI metrics measured at either MRI1 or MRI2. Specifically, reduced PC1 was associated with overall increases in FA, MD, and AD following inpatient treatment (in the Δ scores), across distributed WM anatomical connectivity networks representing primarily frontal, fronto-parietal, and cortico-limbic tracts (encompassing the genu and body of the corpus callosum, fornix, anterior and posterior corona radiata, and superior longitudinal fasciculus for ΔFA and in the superior longitudinal fasciculus and superior corona radiata for ΔMD; correlations with ΔAD were observed in similar regions as well as more posterior cortical tracts). No significant correlation was found with ΔRD. That is, the lower the PC1 score, the higher the FA (and MD and AD) increases with treatment. Follow-up analyses within HC and iHUD groups revealed that the slope differences were driven by significant negative correlations within iHUD, and null results in the HC group.

**Figure 1:**
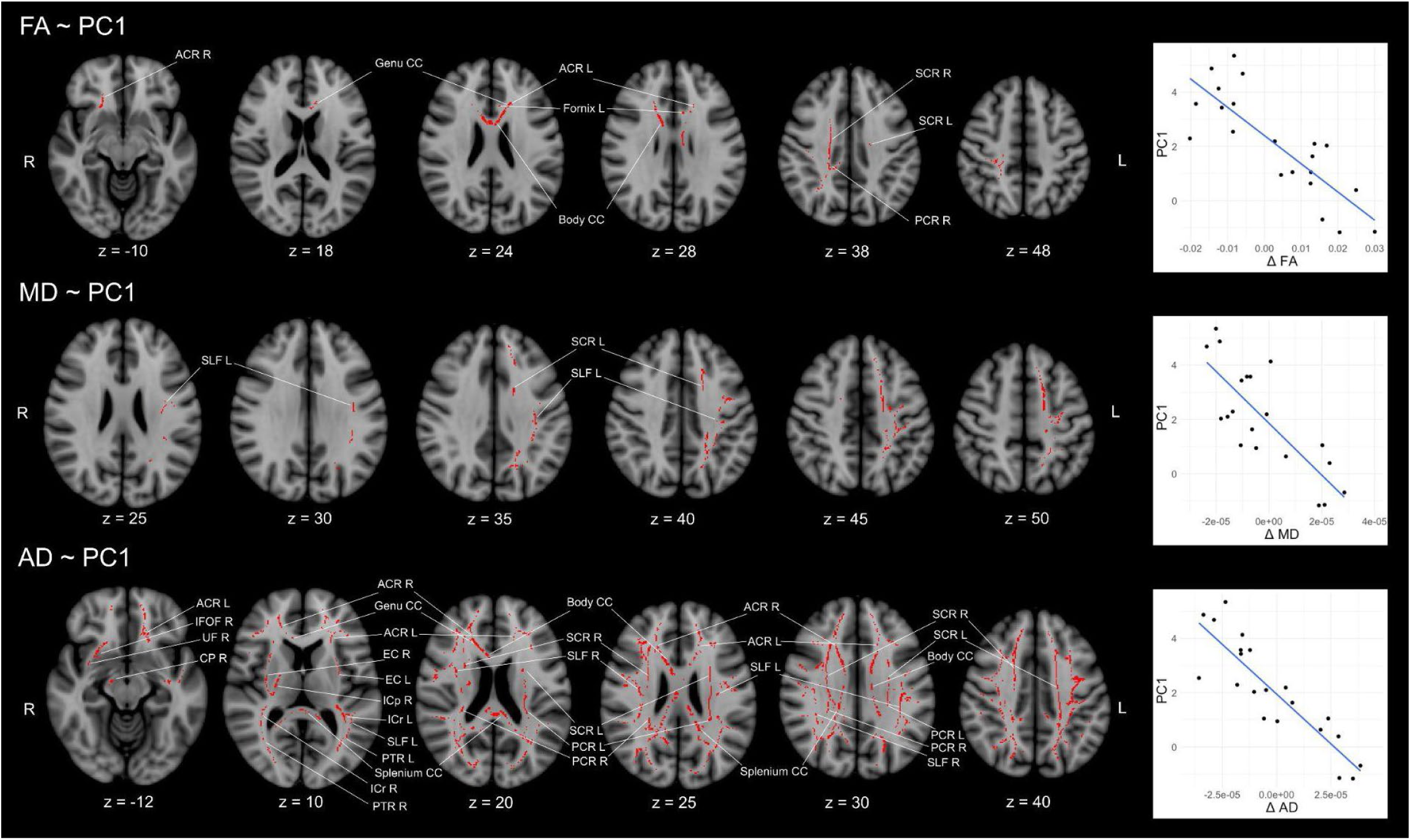
Cytokine PC1 score correlations with Δ (MRI2-MRI1) DTI metrics. Binarized tracts that overlay the significant correlations between the PC1 score (of the 19 cytokines that showed group differences) and the changes in the DTI metrics (FA, MD, AD; non-significant with RD) (axial images); the negative correlations within the iHUD group are shown on the right (examining all significant voxels). Abbreviations: CC: corpus callosum; ACR and PCR anterior and posterior corona radiata respectively; SLF: superior longitudinal fasciculus; PTR: posterior thalamic radiations; IFOF: inferior fronto-occipital fasciculus; UF: uncinate fasciculus; IC and EC: internal and external capsules, respectively); CP: midbrain cerebral peduncles.

As a follow-up, we also examined levels of the cytokine from each canonical family with the greatest PC1 loading for their correlations with ΔDTI metrics (Figures 2-4; see also Table S1 and Figure S1). The results indicated that, among these representative targets, levels of OSM were negatively correlated with ΔFA and ΔAD (Fig. 2), while levels of CSF1 were negatively correlated with ΔMD and ΔAD (Fig. 3), and levels of MP3/CCL7 were negatively correlated with ΔAD (Fig. 4). These correlations occurred over largely overlapping regions for most of the aforementioned cytokines.

**Figure 2:**
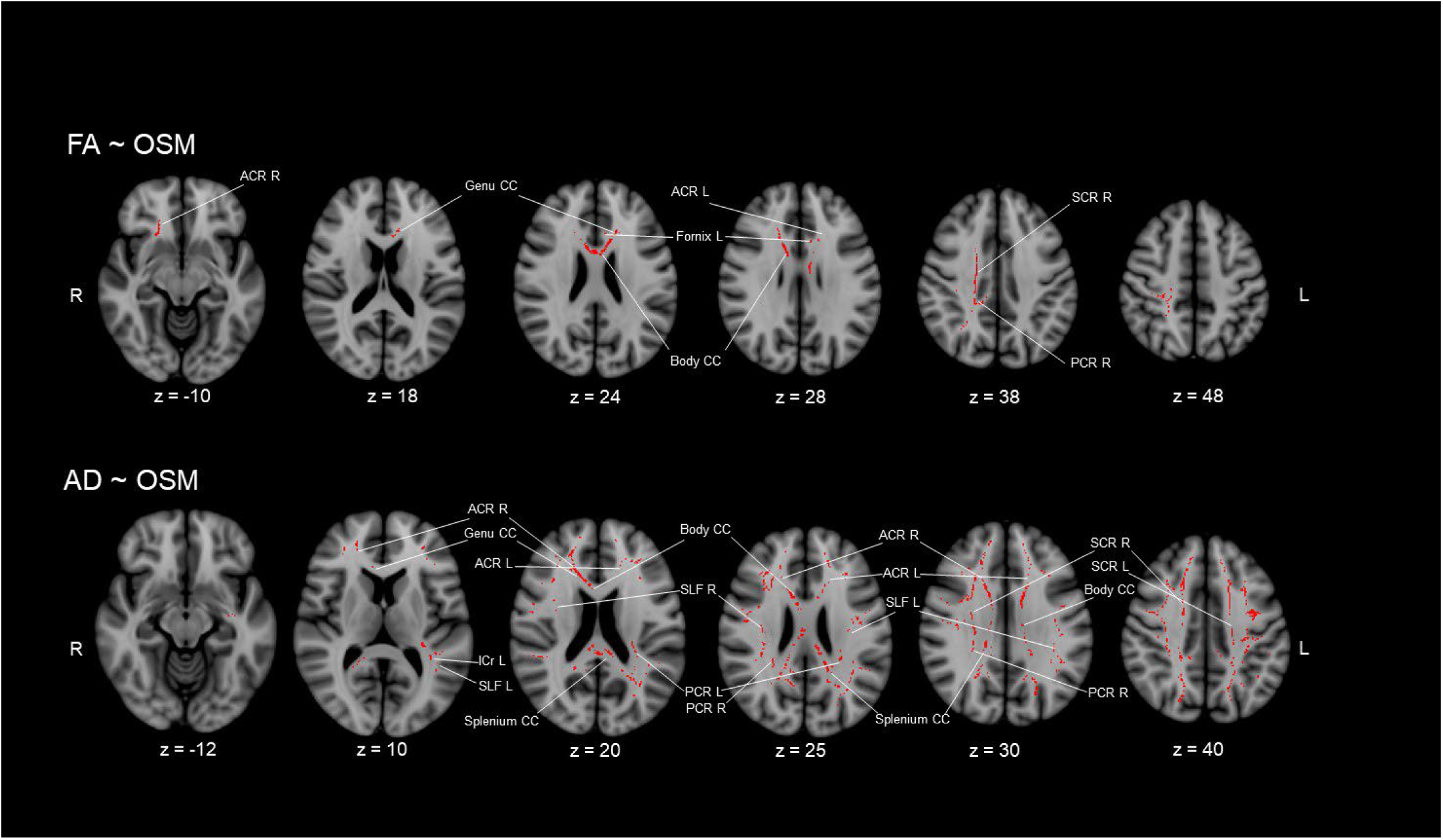
Correlation of oncostatin M (OSM) levels with Δ(i.e., MRI2-MRI1) DTI metrics within the iHUD group. Binarized tracts for within-iHUD negative correlations with serum OSM levels and ΔFA (top row), ΔAD (bottom row). Other details as in Fig. 1.

**Figure 3:**
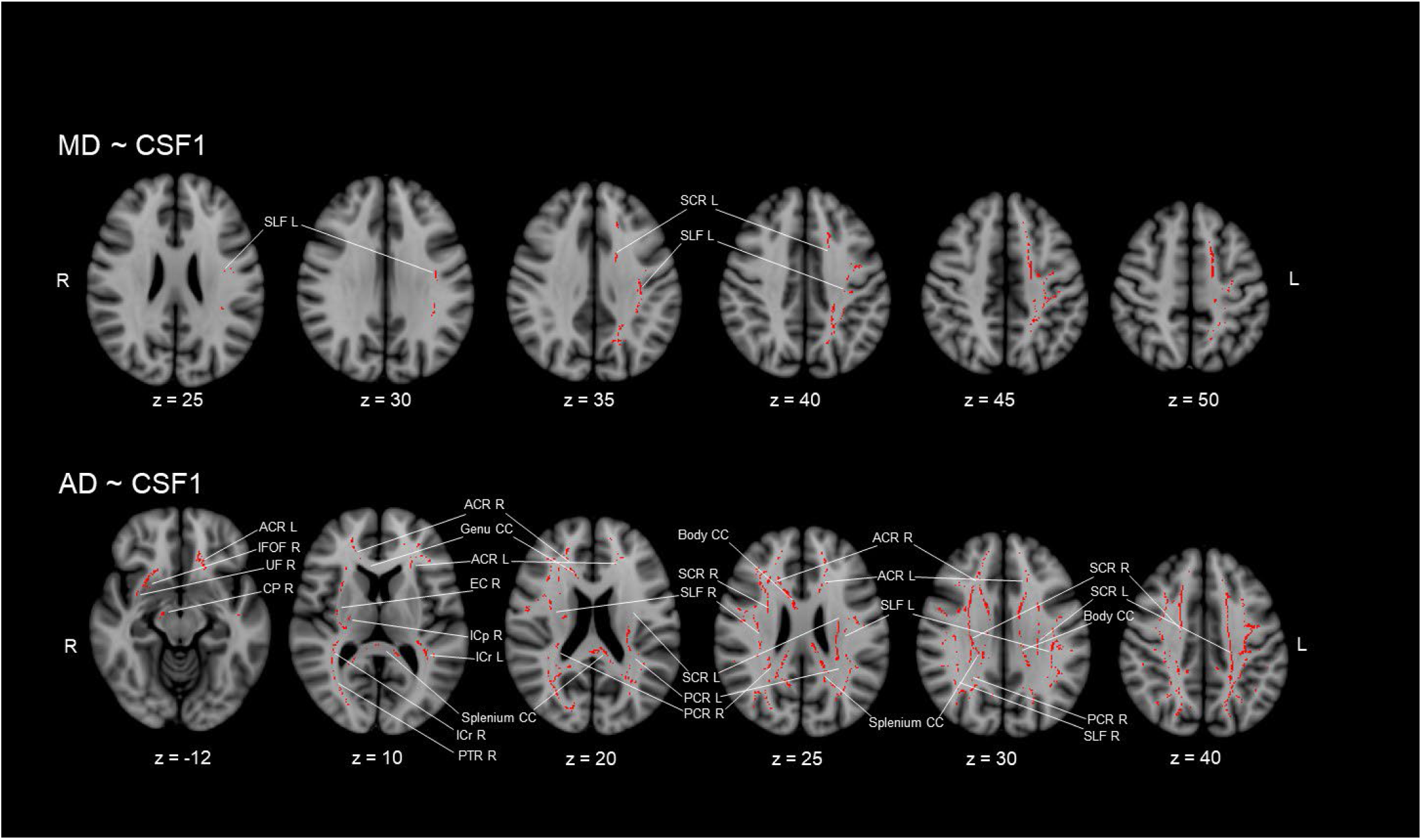
Correlation of colony-stimulating factor levels (CFS1) with Δ(i.e., MRI2-MRI1) DTI metrics within the iHUD group. Binarized tracts for within-iHUD negative correlations with serum CSF1 levels and ΔMD (top row), ΔAD (bottom row). Other details as in Fig. 1.

**Figure 4:**
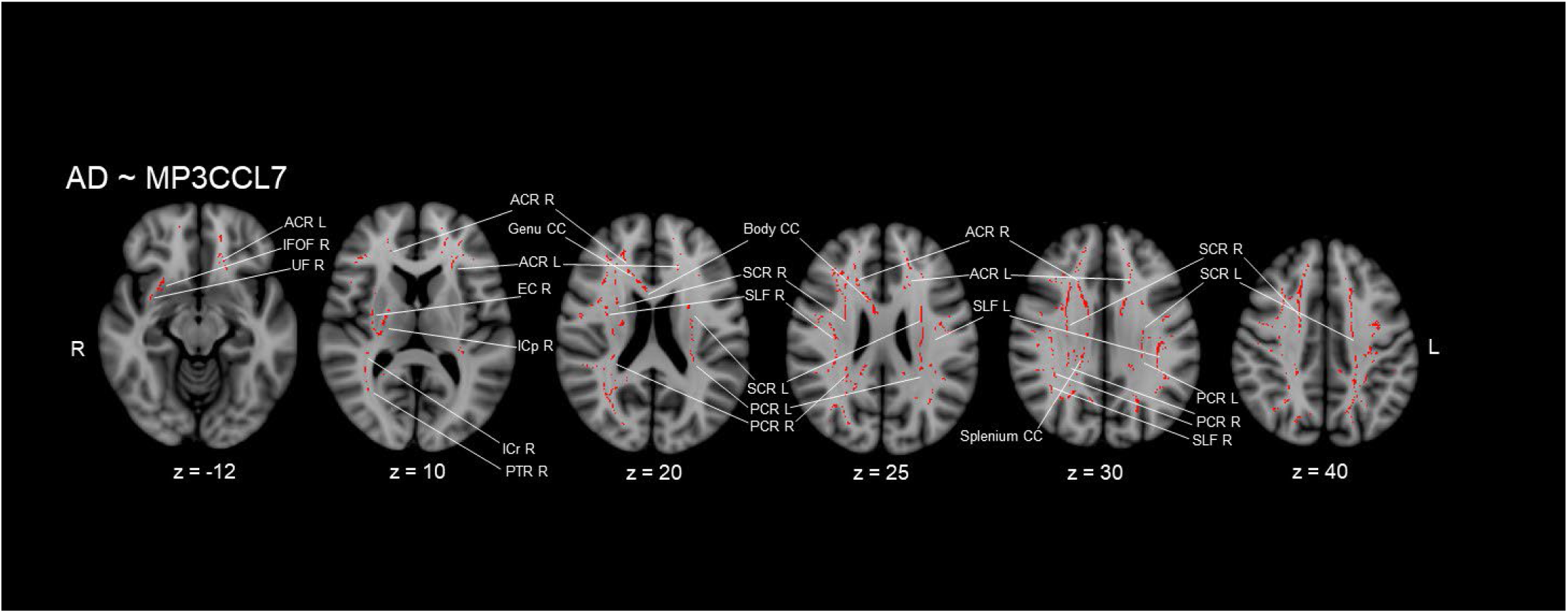
Correlation of monocyte-chemotactic protein 3 levels (also known as chemokine C-C motif ligand 7; CCL7) with Δ(i.e., MRI2-MRI1) DTI metrics within the iHUD group. Binarized tracts for within-iHUD negative correlations with serum MP3 / CCL7 levels and ΔAD are shown. Other details as in Fig. 1.

### Change in cue-induced ratings (MRI2-MRI1)

We first examined whether change in cue-induced ratings of craving, arousal and drug valence were correlated with the overall cytokine PC1 score. Only cue-induced arousal had a significant negative correlation with PC1 scores (Pearson r=-0.46; p-uncorrected=0.034, does not survive FDR correction). However, we found that change in some cue-induced ratings was negatively correlated with levels of specific cytokines. Thus, change in cue-induced cravings was negatively correlated with levels of CCL19 (Pearson r=-0.67; p-uncorrected=0.001; FDR corrected p=0.019; Figure 5A), such that a decrease in cue-induced craving in MRI2 - MRI1 was correlated with greater CCL19 levels. Also, change in cue-induced arousal was negatively correlated with levels of CCL2 (Pearson r=-0.68; p-uncorrected=0.001; FDR corrected p=0.011; Figure 5B), such that a decrease in cue-induced arousal in MRI2 - MRI1 was correlated with greater CCL2 levels. None of the correlations with changes in cue-induced valence ratings survived FDR correction (full data are in Supplement Table S2). Neither PC1 scores, nor levels of any of these 19 cytokines were correlated with cue-induced craving, arousal or valence measured separately at either MRI1 or MRI2, after FDR correction (not shown).

**Figure 5:**
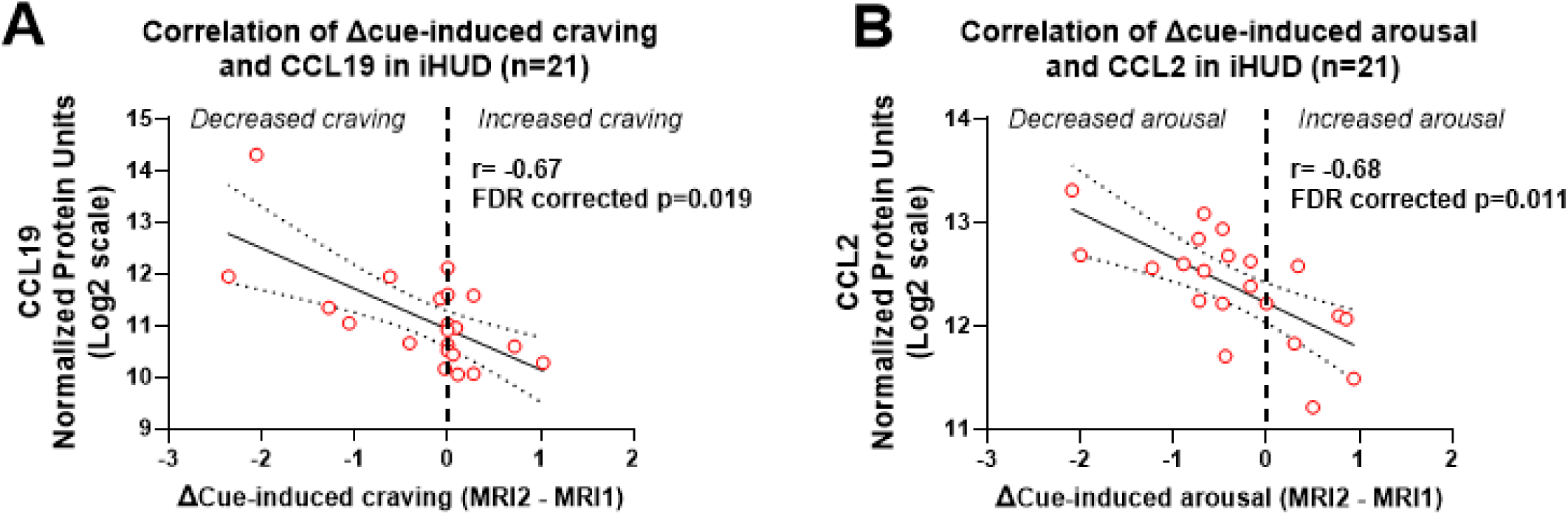
Correlations of changes in cue-induced craving or arousal (MRI2 - MRI1) with specific cytokines in iHUD, measured at MRI2. Pearson correlation is shown, with linear regression and 95%CI bands. **Panel A** shows change in cue-induced craving and CCL19 levels. **Panel B** shows change in cue-induced arousal and CCL2 levels.

## Discussion

The goal of this study was to test the association between cytokine / inflammation blood biomarkers and change in WM microarchitecture, as associated with drug cue-induced responses, following 14 weeks of inpatient MAT in iHUD. The main finding of this study is that a recently developed multi-target cytokine biomarker score (14) was correlated with ΔDTI metrics in several WM tracts. To our knowledge, this is the first study to examine blood levels of cytokines as biomarkers of WM recovery and associated clinical symptoms in iHUD, further expanding knowledge of these blood markers as predictors and mechanisms in psychiatric diseases overall (50,51).

### Serum levels of specific cytokines are correlated with WM change with treatment

We recently reported that these iHUD in MAT had robust dysregulation of serum cytokine levels (14); this new study now shows that the magnitude cytokine elevations/reductions (examined together as a multi-target PCA score) was correlated with WM recovery (ΔDTI) following inpatient treatment. Thus, the cytokine PC1 score was negatively correlated with DTI recovery in iHUD (increases in FA, AD and MD correlated with reductions of cytokines PC1 score) even with a stringent whole-brain approach (49,52), in anterior, posterior, and superior corona radiata, and genu of corpus callosum (Figure 1). In a follow-up (Figures 2-4), we found that several representative cytokines from different canonical families included in the PCA (i.e., IL6 family member OSM, the growth factor CSF1 and the chemokine CCL7) largely recapitulated the above correlations with DTI metrics observed in the overall cytokine biomarker score. Changes in WM microstructure in these and other tracts have been previously associated with neurocognitive dysfunction and disease severity in iHUD (28,53,54) but the mechanisms underlying WM impairment or recovery are not understood. This is the first study in HUD that shows WM and cytokine relationships; some prior clinical studies in other diseases (e.g., major depression) have detected correlations between serum cytokine levels and DTI measures (55,56). This study cannot address causality of serum cytokines and changes in WM microstructure. However, some ancillary evidence may guide future studies. First, there is converging evidence that CSF1 and OSM mediate glial functions including myelination (57–60). Also, the transcription of CSF1 or OSM genes (or that of their cognate receptors) was elevated in post-mortem ventral midbrain of iHUD compared to controls (25). Second, the TNF family cytokine TNFRSF9 (also known as 4-1BB or CD137) is known to mediate oligodendrocyte apoptosis (61,62). Third, the chemokine CCL7 is a chemoattractant for macrophages and can promote neuroinflammation after trauma (63). While little is known on the role of CCL7 in addiction *per se*, forebrain mRNA levels OF CCL7 are increased after acute injection of cocaine in mice and should be further studied in relation to opioid exposure (64). Intriguingly, expression of one of the cognate receptors for CCL7 (CCR10), was upregulated in post-mortem ventral midbrain oligodendrocytes of iHUD versus controls (25). Stem cell factor (65) has multiple effects on cellular lineage (66), it mediates neuro-glial function (67), and is upregulated during inflammatory states (68). Therefore, future studies should examine whether specific cytokines working individually or in networks, are causally involved in changes in WM microstructure during MAT.

### Change in cue-induced ratings (MRI2- MRI1) relate to cytokine levels in the iHUD, at the time of MRI2

We also found that change of cue-induced subjective ratings in the iHUD (i.e., MRI2 - MRI1) were negatively correlated with serum levels of specific cytokines, demonstrating these immune markers may be potentially used as biomarkers of addiction symptoms. Thus, there was a negative correlation between change in cue-induced craving and levels of the chemokine CCL19 (Figure 3, Panel A). Of potential relevance, CCL19 is the ligand for chemokine receptor CCR7 (69), and has been shown to cause heterologous desensitization of MOR *in vitro* (*70*). There was also a negative correlation between change in cue-induced arousal and levels of CCL2 (Figure 3, Panel B) the ligand for chemokine receptors CCR2 and CCR4 (71).

Furthermore, a CCL2 - induced mechanism has recently been implicated in development of opioid withdrawal in a rodent model (16), and increased CCL2 blood levels have been causally associated with decreased FA in humans (72). However, as we recently reported in these same participants, serum levels of both CCL19 and CCL2 were *greater* in iHUD versus HC at the MRI2 stage (14). While these findings are therefore apparently counter-intuitive, two considerations may guide follow-ups: First, it is unknown whether the timeline of specific cytokine serum levels is linear across MAT trajectory. Second, for simplicity, we only examined four representative members of canonical cytokine families for their correlations to ΔDTI measures. It is known that cytokines work in complex functional networks (9); therefore, other targets not examined here could more fully show how patterns of cytokine dysregulation in the iHUD underlie these findings. Overall, future multi-modal longitudinal studies should examine larger serum cytokine networks, together with cue-induced craving/arousal and DTI measures in iHUD.

### Limitations and methodological considerations

First, while it is known that peripheral and central cytokine/immune systems can be functionally connected (16,73), and several of the cytokines examined herein are involved in WM and glial homeostasis, this study cannot address the causality of levels of serum cytokines on changes in WM microstructure, or on subjective ratings such as craving (74–76). However, chronic morphine self-administration can cause robust changes in brain DTI metrics in rats (77). Future studies should therefore examine whether immune and inflammatory changes in iHUD (8,14,41) could potentially contribute to the long-term changes in WM microstructure. Overall, future studies should examine the potential for causal relationships between changes in peripheral serum cytokines and DTI metrics in iHUD, as well as craving and arousal. Second, as a means of dimensionality reduction, we focused here on the 19 cytokines in this multiplex assay that differed significantly between iHUD and HC (14), and larger longitudinal studies could examine a broader array of cytokines that may be important biomarkers for WM microstructure, at different stages in OUD trajectory, such as relapse. Third, interpretation of DTI metrics such as FA can be complicated due to “crossing fibers” or other heterogeneities (78). Of relevance, callosal tracts which among others have shown correlations here, have predominantly parallel fiber organization, and are relatively less vulnerable to such complications (78). Lastly, this study only examined cytokine biomarkers at the MRI2 timepoint; future larger studies should focus on parallel longitudinal analyses of both serum cytokines and DTI metrics.

## Summary and conclusions

This is the first study to examine serum cytokine multiplex levels as biomarkers of longitudinal recovery of WM integrity in iHUD in standard of care MAT. We found that recovery of WM microstructure in specific tracts across 14 weeks in MAT was negatively correlated with cytokine PC1 scores, based on 19 targets which differed significantly in iHUD vs HC (14). This result was also observed when we examined representative cytokines in the PCA (oncostatin M, CSF1 and CCL7) thus implicating specific molecular targets in this phenomenon. Furthermore, levels of other cytokines (i.e., CCL19 and CCL2) were negatively correlated with change in cue-induced craving or arousal across the two timepoints. Overall, only a few prior studies have examined whether pharmacological manipulation of immune and inflammatory processes can modulate MOR-agonist -induced effects in humans (79–83), and none of these studies focused on specific cytokine systems. Thus, the mechanistic bases of dysregulation of cytokine networks and immune disturbance in HUD and WM status are largely unknown (5,8,84–86), and should be the focus of future studies.

Here include future use for therapeutic purposes. Future larger longitudinal studies should examine whether these, or other specific cytokine targets are mechanistically involved in recovery of WM microstructure in iHUD during MAT, and may underlie in part clinical improvement, as well as their diagnostic and prognostic value as biomarkers (87).

## Supporting information

Supplementary Results

## Data Availability

Data in the present work will be available upon reasonable request, after consultation with Institutional Review Board.

## Abbreviations

AD: axial diffusivity
BMI: Body mass index
CCL2: C-C motif chemokine ligand 2
CCL7: C-C motif chemokine ligand 7
CSF1: colony-stimulating factor 1
ΔDTI: change in DTI metrics across longitudinal scans (i.e., MRI2 - MRI1)
DTI: diffusion tensor imaging
FA: Fractional anisotropy
FDR: False discovery rate
HC: Healthy controls
HUD: Heroin use disorder
iHUD: Individuals with heroin use disorder
IL: interleukin
MOR: Mu-opioid receptors
MRI1 or MRI2: First and second longitudinal scans (separated by ≈14 weeks)
NPX: Normalized protein expression (relative quantification from *Olink* assay; log2 units)
OSM: Oncostatin M
PC1 scores: Principal component 1 scores
PCA: Principal component analysis
RD: radial diffusivity

