## Supplementary Results for "Peripheral blood cytokines as markers of longitudinal recovery in white matter microstructure following inpatient treatment for opioid use disorders"

**Supplement**

| **Table S1: Cytokines with significantly different serum levels in iHUD vs. HC (variables in the PCA)** | | | |
|---|---|---|---|
| **Canonical cytokine family** | | | |
| **Growth factor family** | **Tumor necrosis factor (TNF) family** | **Interleukin (IL) family** | **Chemokine family** |
| ***Macrophage-colony Stimulating Factor (CSF1)**** *(1)* | ***TNF receptor superfamily member 9 (TNFRSF9)**** *(2,3)* | ***Oncostatin M (OSM)**** *(4)* | ***C-C motif chemokine ligand 7 (CCL7)**** *(5,6)* |
| Hepatocyte growth factor (HGF) (7) | TNF-related apoptosis-inducing ligand (TRAIL) (8) | IL15 receptor subunit alpha (IL15RA) (9) | C-X-C motif chemokine ligand 9 (CXCL9) (10,11) |
| Transforming growth factor-alpha (TGF-alpha) (12) | TNF ligand superfamily member 14 (TNFSF14) (13,14) | IL18 (15) | C-C motif chemokine ligand 19 (CCL19) ((16) |
| Stem cell factor (SCF)** (17) | TNF receptor superfamily member 5 (TNR5 / CD40) (18) | IL10 receptor subunit beta (IL10RB) (19) | C-C motif chemokine ligand 2 (CCL2) (20) |
| | | IL6 (21–23) | C-C motif chemokine** ligand 28 (CCL28) (24) |
| | | IL7** (25–27) | |

*The cytokine from each family with the PC1 loading of greatest magnitude is in ***bold italics*** (see Fig. S1).

**Only three of the targets were HC>iHUD, targets: SCF, IL7 and CCL28. The remaining sixteen targets were iHUD>HC (28).

**Figure S1. Panel A:** Violin plot of PC1 scores for participants in the two groups (n=24 HC versus n=21 iHUD), examined with a Mann-Whitney test. These PC1 scores are based on the 19 cytokine targets shown in panel B. Data are re-plotted from (28). Panel B: Loadings for principal component analysis 1 (PC1) based on serum levels of the 19 cytokines that differed significantly between iHUD and HC, after false discovery rate (FDR) correction. The target in each canonical cytokine family with the PC1 loading of greater magnitude is in filled squares (i.e., CSF1, TNFRSF9, OSM and CCL7); these targets were examined for their correlation with white matter microstructure metrics (main Results). Targets in each family with PC1 loadings of smaller magnitude are in open squares.

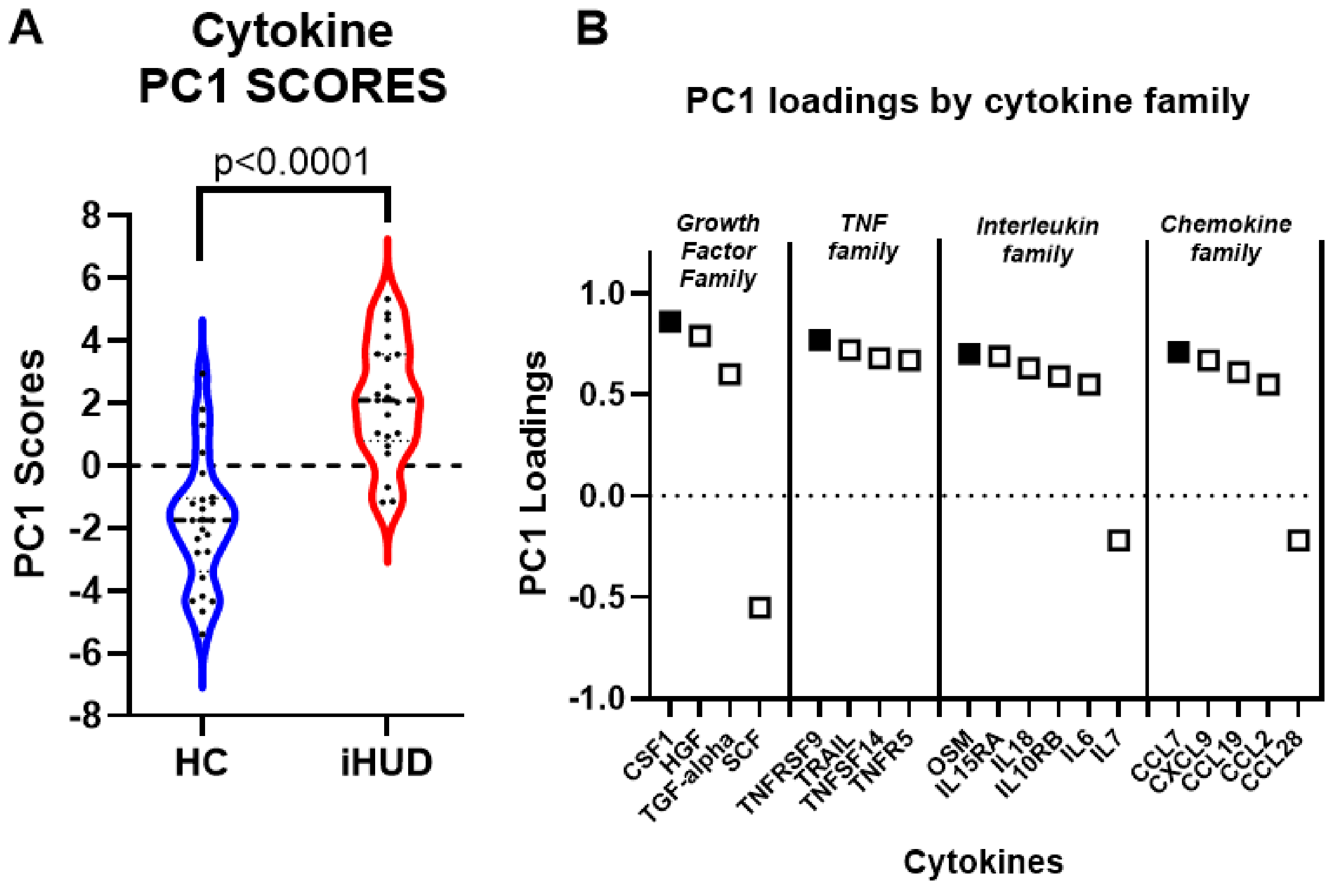

**Figure S2:** Violin plots of the cytokines from each canonical family with the highest PC1 loadings (see Fig. S1; Panel B). Relative quantification of serum levels of CSF1, TNFRSF9, OSM and CCL7 is shown in Panels A-D, respectively. Mann-Whitney tests are shown uncorrected and after FDR correction. Data are re-plotted from (28).

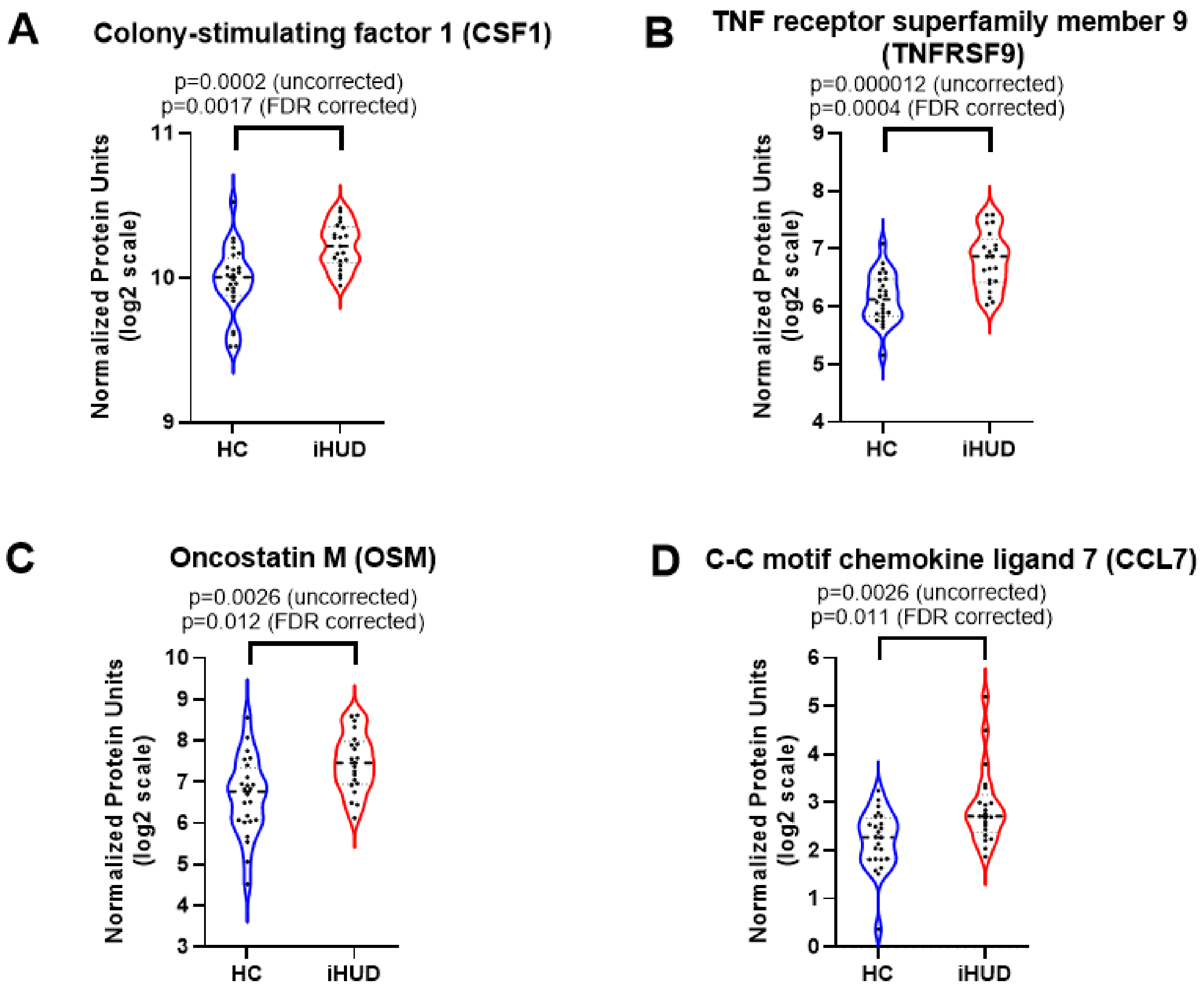

| Table S2: Correlation of individual cytokine levels at MRI2 with Δ in cue-induced ratings (i.e., MRI2 - MRI1) | | | | | | | | | |
|---|---|---|---|---|---|---|---|---|---|
| **Cytokine** | **Cue-induced craving** | | | **Cue-induced arousal** | | | **Cue-induced drug valence** | | |
| | **r[a]** | **p-value[a]** | **FDR[b] p** | **r** | **p-value** | **FDR p** | **r** | **p-value** | **FDR p** |
| CCL7 | -0.13 | 0.560 | 0.999 | -0.23 | 0.315 | 0.977 | -0.081 | 0.727 | 1.000 |
| IL7 | 0.23 | 0.308 | 0.992 | 0.02 | 0.928 | 0.999 | -0.168 | 0.468 | 1.000 |
| IL6 | -0.04 | 0.859 | 1.000 | -0.09 | 0.686 | 0.999 | 0.063 | 0.785 | 1.000 |
| CCL2 | -0.44 | 0.046 | 0.532 | **-0.68** | **0.001** | **0.011** | -0.471 | 0.031 | 0.450 |
| TRAIL | -0.20 | 0.387 | 0.996 | -0.39 | 0.080 | 0.633 | 0.133 | 0.566 | 1.000 |
| CXCL9 | -0.50 | 0.022 | 0.316 | -0.48 | 0.029 | 0.413 | -0.091 | 0.696 | 1.000 |
| OSM | -0.03 | 0.909 | 1.000 | -0.29 | 0.200 | 0.914 | -0.097 | 0.676 | 1.000 |
| SCF | 0.03 | 0.889 | 1.000 | 0.06 | 0.790 | 0.999 | -0.110 | 0.636 | 1.000 |
| IL18 | 0.13 | 0.589 | 0.999 | 0.12 | 0.619 | 0.999 | 0.087 | 0.709 | 1.000 |
| TGFa | -0.01 | 0.964 | 1.000 | -0.07 | 0.753 | 0.999 | -0.130 | 0.576 | 1.000 |
| TNFSF14 | 0.02 | 0.939 | 1.000 | -0.21 | 0.362 | 0.983 | -0.253 | 0.268 | 0.996 |
| CCL19 | **-0.67** | **0.001** | **0.019** | -0.42 | 0.061 | 0.573 | -0.124 | 0.592 | 1.000 |
| IL.15RA | -0.44 | 0.047 | 0.532 | -0.47 | 0.033 | 0.434 | -0.011 | 0.961 | 1.000 |
| IL10RB | -0.26 | 0.251 | 0.982 | -0.06 | 0.782 | 0.999 | -0.099 | 0.670 | 1.000 |

| *Table S2; continued* | | | | | | | | | |
|---|---|---|---|---|---|---|---|---|---|
| **Cytokine** | **Cue-induced craving** | | | **Cue-induced arousal** | | | **Cue-induced drug valence** | | |
| | **r[a]** | **p-value[a]** | **FDR[b] p** | **r** | **p-value** | **FDR p** | **r** | **p-value** | **FDR p** |
| HGF | -0.18 | 0.442 | 0.997 | -0.43 | 0.052 | 0.573 | -0.161 | 0.487 | 1.000 |
| CCL28 | -0.51 | 0.018 | 0.280 | -0.43 | 0.054 | 0.573 | -0.130 | 0.576 | 1.000 |
| CD40 | -0.12 | 0.598 | 0.999 | -0.43 | 0.054 | 0.573 | -0.082 | 0.723 | 1.000 |
| TNFRSF9 | -0.06 | 0.810 | 1.000 | -0.04 | 0.856 | 0.999 | -0.117 | 0.614 | 1.000 |
| CSF1 | -0.21 | 0.368 | 0.996 | -0.17 | 0.459 | 0.993 | -0.080 | 0.729 | 1.000 |

[a]Pearson r
[b]Uncorrected p-value
[c]False Discovery Rate - corrected p; **values surviving correction are in bold**

arthritis. *Clin Exp Rheumatol* 40: 1393–1402.

12. Singh B, Coffey RJ (2014): From wavy hair to naked proteins: the role of transforming growth factor alpha in health and disease. *Semin Cell Dev Biol* 28: 12–21.
13. Albright AV, González-Scarano F (2004): Microarray analysis of activated mixed glial (microglia) and monocyte-derived macrophage gene expression. *J Neuroimmunol* 157: 27–38.
14. Mauri DN, Ebner R, Montgomery RI, Kochel KD, Cheung TC, Yu GL, *et al.* (1998): LIGHT, a new member of the TNF superfamily, and lymphotoxin alpha are ligands for herpesvirus entry mediator. *Immunity* 8: 21–30.
15. Kaplanski G (2018): Interleukin-18: Biological properties and role in disease pathogenesis. *Immunol Rev* 281: 138–153.
16. Nilsson IAK, Millischer V, Göteson A, Hübel C, Thornton LM, Bulik CM, *et al.* (2020): Aberrant inflammatory profile in acute but not recovered anorexia nervosa. *Brain Behav Immun* 88: 718–724.
17. Piao C-S, Li B, Zhang L-J, Zhao L-R (2012): Stem cell factor and granulocyte colony-stimulating factor promote neuronal lineage commitment of neural stem cells. *Differentiation* 83: 17–25.
18. Schönbeck U, Libby P (2001): The CD40/CD154 receptor/ligand dyad. *Cell Mol Life Sci* 58: 4–43.
19. Kotenko SV, Krause CD, Izotova LS, Pollack BP, Wu W, Pestka S (1997): Identification and functional characterization of a second chain of the interleukin-10 receptor complex. *EMBO J* 16: 5894–5903.
20. Zhu Y, Yan P, Wang R, Lai J, Tang H, Xiao X, *et al.* (2023): Opioid-induced fragile-like regulatory T cells contribute to withdrawal. *Cell* 186: 591–606.e23.
21. Tanaka T, Narazaki M, Kishimoto T (2014): IL-6 in inflammation, immunity, and disease. *Cold Spring Harb Perspect Biol* 6: a016295.
22. Re GF, Jia J, Xu Y, Zhang Z, Xie ZR, Kong D, *et al.* (2022): Dynamics and correlations in multiplex immune profiling reveal persistent immune inflammation in male drug users after withdrawal. *Int Immunopharmacol* 107: 108696.
23. Chan YY, Yang SN, Lin JC, Chang JL, Lin JG, Lo WY (2015): Inflammatory response in heroin addicts undergoing methadone maintenance treatment. *Psychiatry Res* 226: 230–234.
24. Mohan T, Deng L, Wang B-Z (2017): CCL28 chemokine: An anchoring point bridging innate and adaptive

immunity. *Int Immunopharmacol* 51: 165–170.

25. Zhang L, Belkowski JS, Briscoe T, Rogers TJ (2012): Regulation of mu opioid receptor expression in developing T cells. *J Neuroimmune Pharmacol* 7: 835–842.

26. Kumar M, Rainville JR, Williams K, Lile JA, Hodes GE, Vassoler FM, Turner JR (2021): Sexually dimorphic neuroimmune response to chronic opioid treatment and withdrawal. *Neuropharmacology* 186: 108469.

27. Guzik TJ, Nosalski R, Maffia P, Drummond GR (2024): Immune and inflammatory mechanisms in hypertension. *Nat Rev Cardiol* 21: 396–416.

28. Butelman ER, Huang Y, Cathomas F, Gaudreault P-O, Roussos P, Russo SJ, *et al.* (2024): Serum cytokines and inflammatory proteins in individuals with heroin use disorder: potential mechanistically based biomarkers for diagnosis. *Transl Psychiatry* 14: 1–9.
